# Ketotifen for Moderate-to-Severe Uremic Pruritus in Chronic Dialysis Patients: A Prospective Observational Study

**DOI:** 10.64898/2026.07.05.26357308

**Authors:** Kaandeeban Mohanraj, Andrew Deepak Rajiv, Sampathkumar Krishnaswamy

## Abstract

**Background:** Uremic pruritus affects up to 90% of individuals and substantially impairs quality of life, in the group of patients undergoing chronic dialysis. Despite multiple therapeutic options, an optimal and well tolerated treatment remains elusive. Ketotifen, a mast cell stabilizer with antihistaminic properties prevent itch by inhibition of mast cell derived tryptase, which modulates protease-activated receptor-2 (PAR-2) in the cowhage itch pathway.

**Materials and Methods:** In this prospective observational study, 230 chronic dialysis patients were screened, of whom 48 (20.9%) had clinically significant pruritus identified using a structured questionnaire. Twenty-four patients with moderate-to-severe symptoms who were prescribed ketotifen as part of routine clinical care consented to prospective follow-up. Ketotifen was initiated at 1 mg twice daily, with dose escalation to 2 mg twice daily in patients with persistent symptoms according to routine clinical practice. Pruritus severity was assessed using visual (VAS), verbal (VRS), and numerical (NRS) rating scales before and after treatment.

**Results:** After two weeks of initial 1mg therapy, 19 showed significant clinical improvement. Mean scores reduced 77.5 → 27.1 (VAS), 87.5 → 20.8 (VRS), and 74.2 → 25 (NRS) (around 65% reduction with p < 0.001 across all scales). Clinical relief was achieved in 83.3% overall and mild tolerable drowsiness occurred only at the 2mg dose.

**Conclusion:** Ketotifen is a safe, effective and well tolerated option for moderate to severe uremic pruritus in dialysis patients. Larger multicenter studies are warranted to confirm efficacy and optimize dosing.

## Introduction

Uremic pruritus, also known as chronic kidney disease-associated pruritus (CKD-aP), occurs due to the accumulation of uremic toxins in patients with renal failure [1,2]. It is a prevalent complication among patients with chronic kidney disease (CKD) and end-stage kidney disease (ESKD) undergoing dialysis, with reported prevalence ranging from 20% to 90%, and estimates indicating 50–90% prevalence among hemodialysis (HD) patients [3,4]. Meta-analyses suggest similar prevalence rates in hemodialysis (HD) and peritoneal dialysis (PD) patients.

Pruritus severity ranges from mild to severe, significantly impacting patient quality of life globally. Current treatments include serum calcium and phosphate correction, dialysis optimization, topical agents such as paraffin, glycerol, γ-linoleic acid cream, capsaicin, and tacrolimus, as well as systemic therapies like antihistamines, gabapentin, and pregabalin [5,6].

The pathophysiology of CKD-aP is multifactorial. While histamine dependent unmyelinated C fibers contribute to itching, the limited effectiveness of antihistamines suggests involvement of an alternative, non-histaminergic cowhage pathway. Studies show this pathway involves activation of protease-activated receptor 2 (PAR-2) in the skin, transmitting itch via polymodal C neurons [7,8].

Mast cell stabilizers, such as cromolyn sodium and ketotifen, are established treatments for allergic conditions. Ketotifen, an inexpensive oral medication with additional antihistaminic action specifically shows potential therapy for CKD-aP by addressing both histaminergic and non-histaminergic mechanisms implicated in uremic pruritus [9–12].

This study prospectively evaluated the clinical outcomes of routine ketotifen therapy in patients with uremic pruritus undergoing chronic hemodialysis, using validated pruritus severity scales at baseline and after two and four weeks of follow-up. Secondary objectives were to assess the tolerability of ketotifen and changes in patient-reported quality of life, while exploring its potential role as a cost-effective treatment option for moderate-to-severe CKD-associated pruritus in routine clinical practice.

## Materials and Methods

### 1. Study design

This prospective observational study was conducted at the dialysis unit of Meenakshi Mission Hospital and Research Center, Madurai, Tamil Nadu, India, to evaluate the clinical outcomes and safety of routine ketotifen therapy in treating chronic kidney disease-associated pruritus (CKD-aP) among patients on hemodialysis.

### 2. Participants

Twenty-four patients meeting inclusion criteria who were prescribed ketotifen as part of routine clinical care provided informed consent and were enrolled in the study.

### 3. Inclusion criteria

a. Clinically significant pruritus diagnosed as as *CKD-associated pruritus (CKD-aP)* based on: **Onset:** Itching in CKD patients without other identifiable causes; **Frequency and duration:** Itching occurring at least three times over two weeks, multiple times per day, lasting several minutes and causing discomfort; **Chronic pattern:** Recurrent itching for ≥ 6 months, even if intermittent [2]
b. Moderate-to-severe pruritus severity, assessed using standardized scales (VAS, VRS, NRS)
c. Provided written informed consent

### 4. Exclusion criteria

Patients having mild pruritus, Other dermatologic or systemic conditions causing pruritus (e.g., psoriasis, atopic dermatitis, cholestatic liver disease, thyroid dysfunction) or those who declined participation (n=5) were excluded.

### 5. Dialysis procedure

Patients underwent twice-weekly hemodialysis sessions, each lasting between 3 to 5 hours, utilizing standard dialysis machines with bicarbonate dialysate solutions. All procedures followed established clinical protocols for patient monitoring and safety.

### 6. Routine treatment

Patients prescribed oral ketotifen as part of routine clinical care initially received ketotifen 1 mg twice daily for two weeks. Patients with persistent symptoms (n=5) had the dose escalated to 2 mg twice daily as part of routine clinical care [13].

### 7. Outcome assessment

Pruritus severity was evaluated at baseline and post treatment using three validated scales [14]:

a. **Visual Analogue Scale (VAS):** Patients indicated itch severity visually, matched numerically from 0 (no itch) to 10 (worst imaginable itch).
b. **Verbal Rating Scale (VRS):** Categorized pruritus as 0 (no itch), 1 (mild), 2 (moderate), and 3 (severe).
c. **Numerical Rating Scale (NRS):** Scored from 0 (no itch), 5 (moderate itch), to 10 (worst imaginable itch).

For reference, the 14-item **UP-Dial Questionnaire** was used to cross-validate the accuracy of the simplified questionnaire used in this study, ensuring consistency with established chronic kidney disease associated pruritus assessment methods [15].

### 8. Data collection

Data were collected prospectively by the principal investigator, who identified eligible patients, recorded demographic and clinical characteristics, and conducted scheduled follow-up assessments to document changes in pruritus severity and adverse events. Treatment decisions, including ketotifen initiation and dose escalation, were made by the treating nephrologist as part of routine clinical care.

Data collection involved a structured “Pruritus in Dialysis Questionnaire,” completed during dialysis sessions. [Supplementary File 1] Collected data included:

a. **Patient Information:** Age, hospital number, height, weight
b. **Clinical Parameters:** Systolic and diastolic blood pressure
c. **Clinical Signs:** Presence of edema, skin papules, and shiny nails
d. **Medical History:** Etiology of renal disease (diabetes, hypertension, chronic glomerulonephritis, or others), dialysis duration and total sessions
e. **Laboratory Results:** Hemoglobin, total leukocyte count (TC), erythrocyte sedimentation rate (ESR), pre-dialysis blood urea and creatinine levels, blood sugar, HbA1c, calcium, phosphorus, uric acid
f. **Treatment Details:** Use of calcium binder tablets (calcium carbonate, sevelamer, or others) and previous antipruritic drug usage
g. **Sleep Disturbance:** Presence of sleep disturbances due to itching

Assessments were conducted at baseline, at end of two weeks (1 mg BID), and at four weeks for those receiving 2 mg BID.

### 9. Statistical analysis and adverse event monitoring

Data were entered into Microsoft Excel and analyzed using *jamovi* software (Version 2.6). For consistency across scales, all pruritus scores were normalized to a 0 to 100% range. The percentage change from baseline to post treatment was calculated manually for descriptive reporting. Paired t-tests were performed on the normalized (percentage) scores to determine mean within-subject differences baseline and followup. A p-value <0.05 was considered statistically significant.

Participants were monitored for adverse reactions throughout the study. Events were promptly documented by dialysis staff and graded using the *Common Terminology Criteria for Adverse Events (CTCAE), version 5.0*. Mild events were managed symptomatically, while moderate or severe reactions were managed according to routine clinical practice.

### 10. Ethical considerations

Ethical approval was obtained from the Ethics Committee of Meenakshi Mission Hospital and Research Center. Written informed consent was obtained from all participants for prospective data collection, and confidentiality was maintained throughout the study in accordance with the Declaration of Helsinki.

## Results

Of the 230 patients screened, 48 (20.9%) had clinically significant pruritus, with 29 (60.4%) experiencing moderate-to-severe symptoms and 19 (39.6%) mild symptoms.

Table 1 shows the Demographic, Clinical, and Laboratory Profile of the 48 patients with clinically significant pruritus.

**Table 1:**
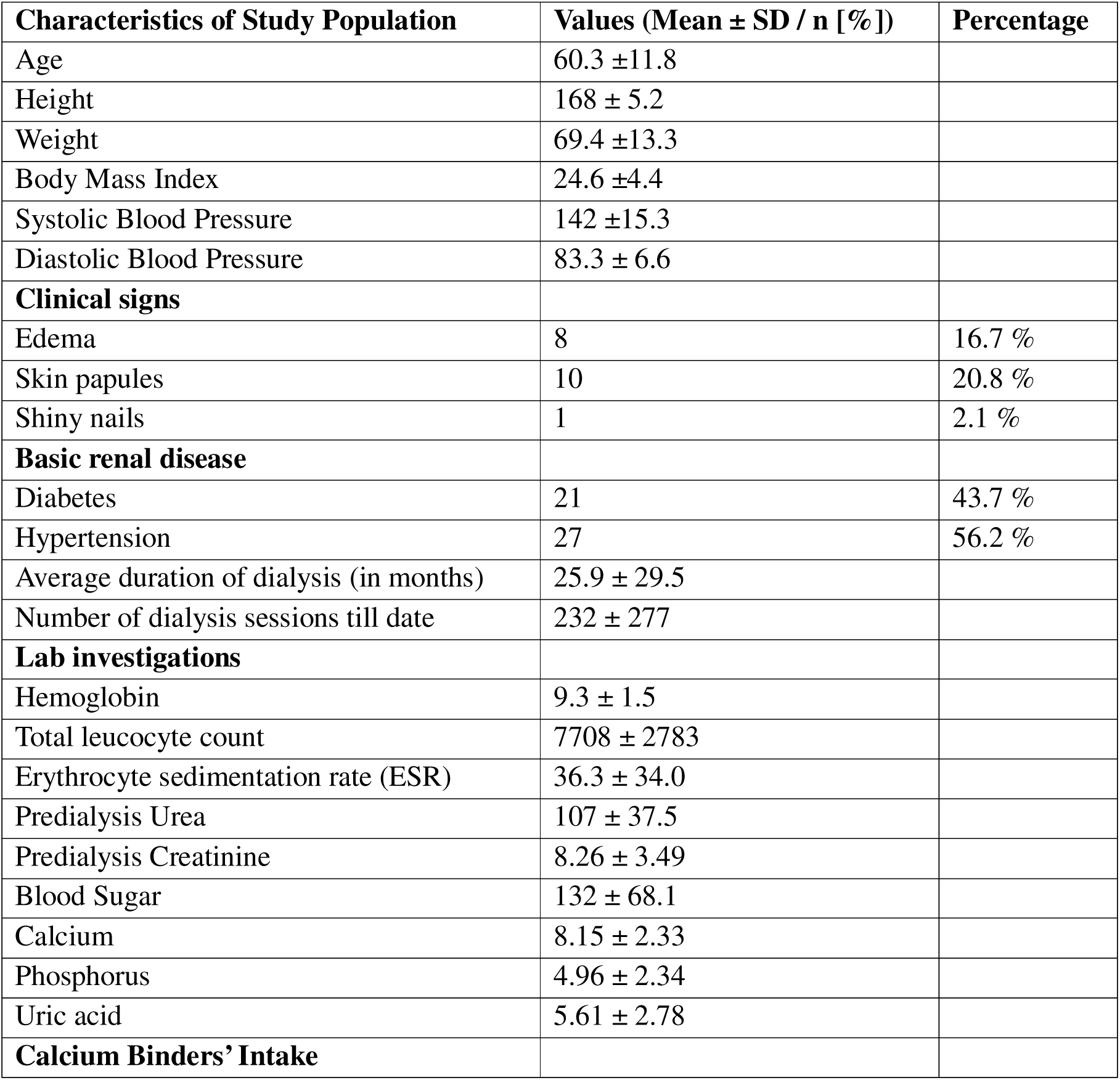
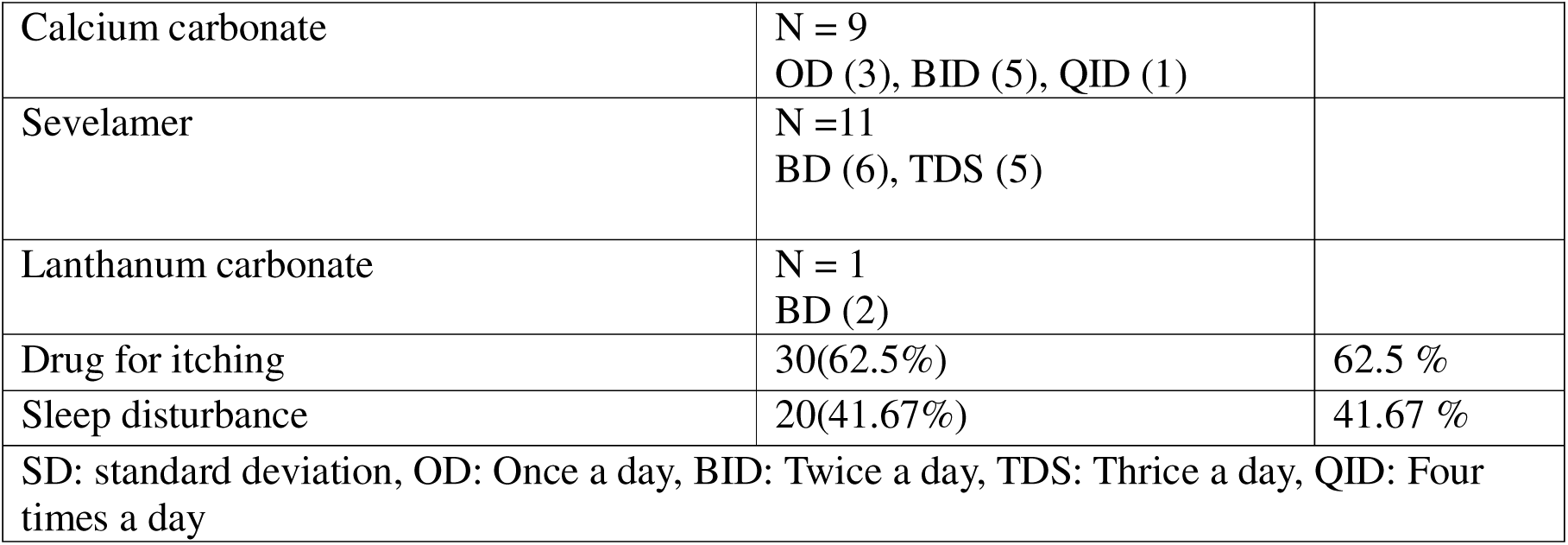
Demographic, Clinical, and Laboratory Profile of the Baseline Uremic Pruritus population (N = 48)

Participants had a mean age of 60.3 ± 11.8 years, predominantly male (87.5%, n=42), with most patients in the age groups 51-60 and 61-70 years. [Figure 1] Mean height, weight, and BMI were 168 ± 5.2 cm, 69.4 ± 13.3 kg, and 24.6 ± 4.4, respectively.

**Figure 1:**
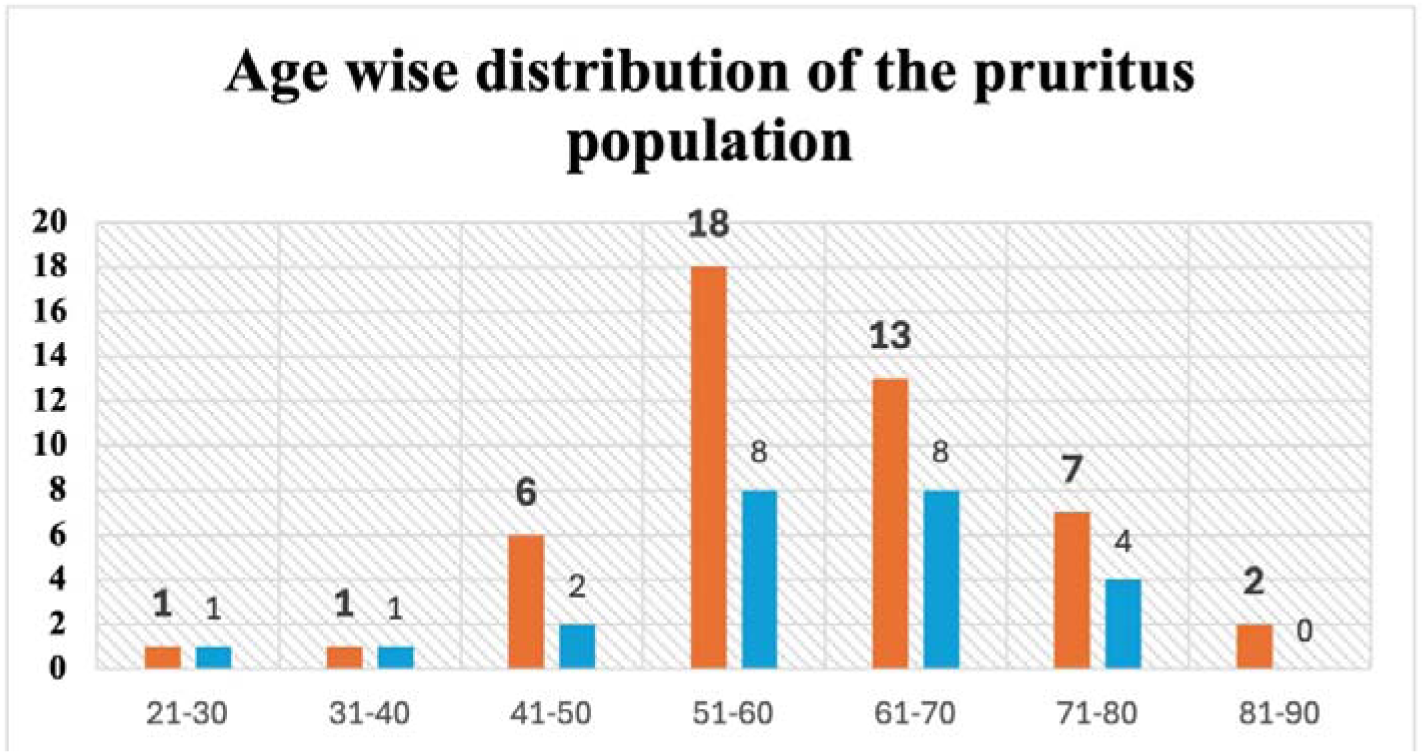
Age-wise distribution of the study population with pruritus and participants enrolled in the prospective observational study. The orange bars show the total number of patients with chronic kidney disease-related pruritus across different age groups. The blue bars indicate patients included in the prospective observational study. Most participants were in the 51- 60- and 61-70-year age groups.

Hypertension was the most common underlying renal disease (56.2%, n=27), followed by diabetes (43.7%, n=21). The mean dialysis duration was 25.9 ± 29.5 months, with a mean of 232 ± 277 dialysis sessions per patient.

Clinically, skin papules were observed in 10 patients (20.8%), pedal edema in 8 (16.7%), and shiny nails in 1 (2.1%). Anti pruritic medications were used by 30 patients (62.5%), and sleep disturbances due to itching were reported by 20 (41.7%).

Lab investigations result of the participants was collected to assess the patients’ health status and potential factors contributing to uremic pruritus.

The hemoglobin level indicates anemia with a level of 9.4 ± 1.5 g/dL. The total white blood cell count is 7708 ± 2783 cells/mm³, while the erythrocyte sedimentation rate (ESR) is 36.3 ± 34.0 mm/hr. Predialysis urea and creatinine levels are elevated at 107 ± 37.5 mg/dL and 8.26 ± 3.5 mg/dL, respectively, confirming the impaired renal function. Blood sugar levels average 132 ± 68.1 mg/dL. Electrolyte reports show serum calcium value of 8.1 ± 2.3 mg/dL, phosphorus of 4.9 ± 2.3 mg/dL, and uric acid of 5.6 ± 2.7 mg/dL.

The baseline mean normalized pruritus scores percentages were 77.5%, 87.5% and 74.2% across VAS, VRS, and NRS scales respectively.

After two weeks of routine ketotifen therapy (1 mg twice daily), 19 of 24 patients (79.2%) showed clinically significant improvement.

Among the five patients who underwent dose escalation to 2 mg twice daily as part of routine clinical care, one patient (20%) showed improvement.

Percentage reductions in pruritus severity were calculated manually using normalized (0–100%) scale values. Visual, verbal, and numerical scores decreased from 77.5 to 9.9, 87.5 to 20.8, and 74.2 to 4.8, respectively, corresponding to an overall improvement of approximately 66 to 69 %. [Figure 2]

**Figure 2:**
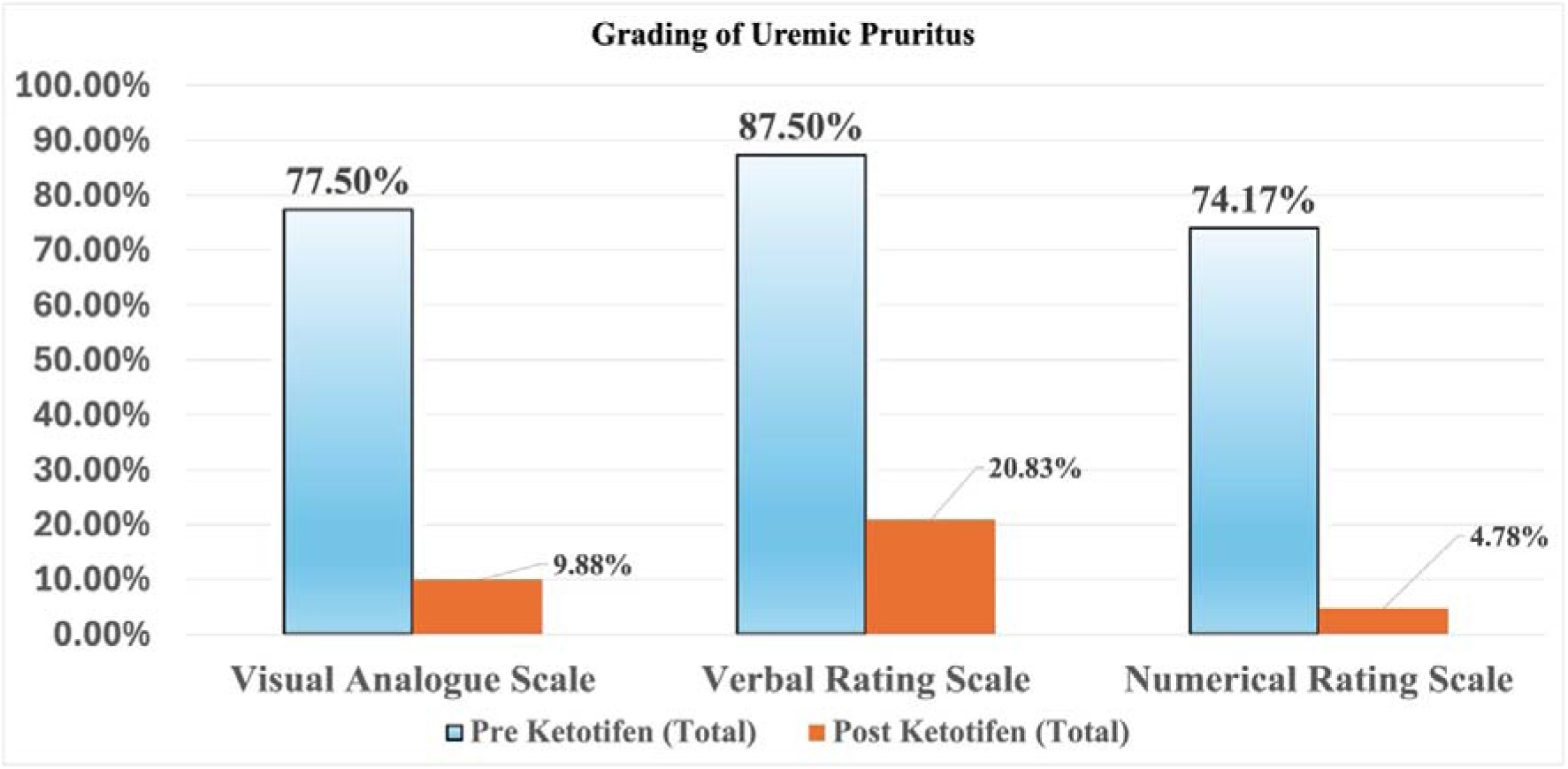
Grading of uremic pruritus before and after routine ketotifen therapy (n = 24). A bar diagram shows the average scores on the visual, verbal, and numerical scales before and during follow-up after routine ketotifen therapy. The percentage reduction in pruritus severity was calculated as 87.25%, 76.19%, and 93.55%, respectively. This indicates a significant improvement in symptoms after treatment.

Overall, 20 of the 24 patients (83.3%) included in the study demonstrated clinically significant relief and patient-reported quality of life improvements. To statistically confirm these findings, paired *t*-tests were performed on the percentage-converted data.

Table 2 shows the paired T test results obtained from statistical analysis (jamovi v2.6)

**Table 2:** Paired Samples t-Test Comparing Baseline and Follow-up Pruritus Scores (N = 24)

| Paired Samples T-Test |  |  |  |  |  |  |  |  |  |  |  |  |  |
| --- | --- | --- | --- | --- | --- | --- | --- | --- | --- | --- | --- | --- | --- |
|  |  |  |  |  |  |  |  | 95%<br>Confiden<br>ce<br>Interval |  |  |  | 95%<br>Confiden<br>ce<br>Interval |  |
|  |  |  | stati<br>stic | df | p | Mean<br>differ<br>ence | SE<br>differ<br>ence | Lo<br>we<br>r | Up<br>per |  | Eff<br>ect<br>Siz<br>e | Lo<br>we<br>r | Up<br>per |
| PRE-VISUA<br>L | POST<br>VISUA<br>L | Stud<br>ent's<br>t | 8.12 | 23<br>.0 | <.0<br>01 | 50.4 | 6.21 | 37.<br>6 | 63.<br>3 | Coh<br>en's<br>d | 1.6<br>6 | 1.0<br>3 | 2.2<br>7 |
| PRE<br>VERBA<br>L | POST<br>VERBA<br>L | Stud<br>ent's<br>t | 8.67 | 23<br>.0 | <.0<br>01 | 58.3 | 6.73 | 44.<br>4 | 72.<br>3 | Coh<br>en's<br>d | 1.7<br>7 | 1.1<br>2 | 2.4<br>1 |
| PRE-NUME<br>RICAL | POST<br>NUME<br>RICAL | Stud<br>ent's<br>t | 8.71 | 23<br>.0 | <.0<br>01 | 49.2 | 5.64 | 37.<br>5 | 60.<br>8 | Coh<br>en's<br>d | 1.7<br>8 | 1.1<br>2 | 2.4<br>2 |
| Note. $H_0: \mu_{\text{Measure 1}} - \mu_{\text{Measure 2}} \neq 0$<br>*df*: degrees of freedom; *SE*: standard error; *CI*: confidence interval; *p*: probability value;<br>*Cohen's d*: effect size. | | | | | | | | | | | | | |
| The jamovi project (2024). *jamovi (Version 2.6)* [Computer Software]. Retrieved from<br>https://www.jamovi.org |  |  |  |  |  |  |  |  |  |  |  |  |  |

The analyses demonstrated significant mean reductions across all scales (VAS: 50.4 ± 6.2, VRS: 58.3 ± 6.7, NRS: 49.2 ± 5.6; *p* < 0.001 for all), with large effect sizes (Cohen’s *d* = 1.66–1.78), demonstrating a significant improvement in pruritus severity during follow-up. [Figure 3]

**Figure 3:**
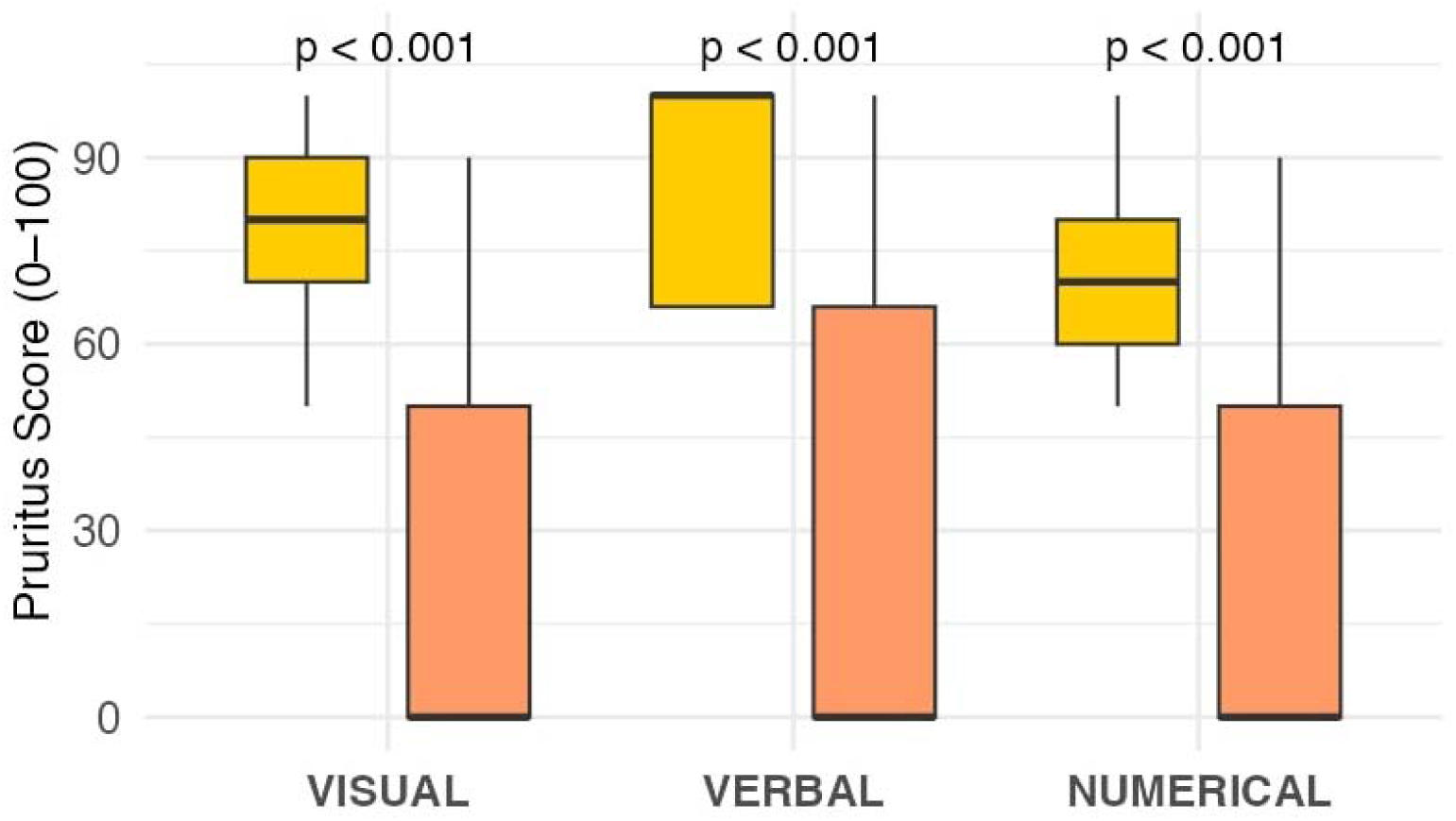
Comparison of baseline and follow-up pruritus scores across visual, verbal, and numerical scales (n = 24). Box-and-whisker plots show the distribution of baseline (yellow) and follow-up (orange) pruritus scores on the three assessment scales. There is a clear reduction after ketotifen therapy, showing significant improvement in symptoms. (p <0.001) Figure generated using The jamovi project (2024). jamovi (Version 2.6) [Computer Software]. Retrieved from https://www.jamovi.org; R Core Team (2024). R: A Language and Environment for Statistical Computing (Version 4.4) [Computer Software]. Retrieved from https://cran.r-project.org (R packages retrieved from CRAN snapshot 2024-08-07).

Four patients (16.7%) did not respond even after dose escalation. Mild, well-tolerated drowsiness with grade 1 on common terminology criteria for adverse events (CTCAE v5.0) occurred in the five patients escalated to 2 mg BID. No severe adverse effects were reported, and importantly none of the participants withdrew from the study due to adverse effects confirming ketotifen’s favorable safety and tolerability profile.

## Discussion

In the subset of patients that did not respond to ketotifen despite dose escalation, several possible factors may underlie in this lack of response. Non-responders were generally older, between 62 and 74 years, compared to the overall group. This suggests that age related pharmacokinetic or sensory differences. There was a higher rate of diabetes among non-responders at 75%, compared to 43.7% in the general group. This points to possible nerve-related or metabolic factors. The length of dialysis varied a lot, indicating different responses to treatment. Non-responders also showed a calcium-phosphorus imbalance, with low calcium and high phosphorus levels. Their elevated ESR, ranging from 50 to 72 mm/hr, suggests ongoing metabolic and inflammatory issues that might reduce the effectiveness of ketotifen.

Historically, antihistamines were the first-line treatments for pruritus, based on the traditional assumption that histamine was the primary mediator of itching. However, trials using hydroxyzine, bilastine, chlorpheniramine, and loratadine consistently shown limited efficacy, often achieving improvement rates less than 50%. [16–19] Randomized controlled trials from Iran and Greece demonstrated relatively modest response rates with antihistamines, indicating that histamine-driven pathways may not be solely responsible for uremic pruritus. These findings have shifted the attention towards role of mast cell stabilizers and neurologically active agents, in the complex pathophysiology underlying uremic pruritus.

Recent research has highlighted greater therapeutic promise in drugs exhibiting mast cell-stabilizing properties. [9–12]

In an Iranian study conducted in 2016, ketotifen (1 mg twice daily) and gabapentin (100 mg daily) yielded comparable improvements of 76.9% versus 88.4% respectively. [12] Furthermore, a recent 2022 Iranian study demonstrated significant improvements in patient-reported quality of life with ketotifen compared to pregabalin, further validating the role of mast cell stabilizers. [11] Topical and oral cromolyn sodium cream has also provided consistent positive outcomes, reinforcing the therapeutic value of mast cell stabilization in chronic pruritus management. [9,10]

Additionally, low-dose doxepin, a tricyclic antidepressant with potent antihistaminic effects, demonstrated a complete relief rate of approximately 58%, although sedation was notably higher. [20]

Gabapentinoids, particularly gabapentin and pregabalin, possess the most robust clinical evidence for managing uremic pruritus demonstrating pruritus severity reductions of up to 85% across multiple trials. Similarly, pregabalin exhibits considerable efficacy, though slightly inferior to gabapentin. Compared to the clinical improvement observed in our study, gabapentin may provide more rapid and pronounced symptom relief but carries risk of CNS side effects such as sedation, dizziness and requires renal dose adjustment.

In contrast, ketotifen offers advantages of hepatic metabolism limits the need for renal dose adjustments, low dialyzability, and minimal CNS-related adverse effects. [20–24]

In this study, the mild sedation induced by ketotifen did not prompt discontinuation, highlighting its favorable safety profile. Thus, the findings of this study suggest that routine ketotifen therapy may represent an attractive, safer, and cost-effective treatment option, particularly suited to resource-limited settings.

Another innovative treatment recently approved for managing CKD-associated pruritus is difelikefalin, a novel kappa-opioid receptor agonist. Clinical trials such as KALM-1 and KALM-2 reported improvement rates of 50.9% and 53.4%, respectively, significantly superior to placebo. Additionally, difelikefalin requires intravenous administration post-dialysis at a dosage of 0.5 μg/kg, which could limit its practicality and accessibility due to higher costs, and limited global availability. In contrast, ketotifen is orally administered, affordable, globally accessible, and pharmacologically suitable for hemodialysis patients [25–31]

Mechanistically, ketotifen exerts effects primarily via mast cell stabilization and inhibition of histamine release, affecting both histaminergic and non-histaminergic (cowhage) itch pathways through tryptase suppression. In comparison, other existing therapies act via distinct points on these itch pathways thus suggesting the potential for combination therapies, where concurrent use of both agents could yield additive or synergistic benefits, warranting future clinical studies. [8]

This study’s limitations must be acknowledged. The relatively small sample size (n=24) restricts the generalizability of our findings to broader dialysis populations. Although the observational design and absence of a placebo-controlled comparison limit causal inference, the improvements observed during follow-up suggest a potential association between routine ketotifen therapy and reduced pruritus severity. The short follow-up duration restricts assessment of long-term efficacy and durability of symptom improvement. Additionally, the potential for observer or measurement bias cannot be completely excluded.

Future research should prioritize larger multicenter prospective comparative studies and randomized placebo-controlled trials with prolonged follow-up to further evaluate the long-term effectiveness and safety of ketotifen, firmly establishing its therapeutic position among current and emerging CKD pruritus treatments.

Furthermore, additional exploration into optimal dosing regimens, patient subgroup responsiveness, and potential combination strategies with gabapentinoids or opioid receptor modulators is warranted to refine and maximize therapeutic outcomes for patients suffering from this debilitating condition.

## Conclusion

Routine ketotifen therapy was associated with clinically meaningful improvement in chronic itching among patients undergoing chronic hemodialysis. The observed improvement may be related to its effects on both histamine-related and non-histamine-related pathways through mast cell stabilization and inhibition of tryptase. At a dose of 1 mg twice daily, ketotifen was affordable, well tolerated, and easy to administer, supporting its potential role as a practical treatment option with global accessibility. Mild sedation was observed only at higher doses. Improving dialysis and skin moisturization may further improve outcomes. While these findings are promising, larger prospective comparative studies and randomized controlled trials are needed to better define the role of ketotifen in the management of CKD-associated pruritus.

## Supporting information

Supplementary file 1. Pruritus in Dialysis Questionnaire

## Author Contributions

**KM:** Conceptualization, study design, data collection, data curation, formal analysis, investigation, methodology, visualization, interpretation of results, writing - original draft, writing - review and editing, and project administration. **ADR:** Investigation, data interpretation, writing—review and editing, and clinical input. **SK:** Conceptualization, supervision, methodology, validation, data interpretation, writing—review and editing, and overall supervision of the study. All authors read and approved the final manuscript.

## Data availability

All data generated or analyzed during this study are included in this manuscript and its supplementary material. Additional de-identified data are available from the corresponding author upon reasonable request.

## Acknowledgment

The authors sincerely thank all participants and the dialysis unit staff for their cooperation and support during the study.

This work was presented as an *e-poster* at the **Royal College of Physicians Medicine 2025 Conference**, and the abstract was published as:

Mohanraj K, Rajiv AD, Krishnaswamy S. *Relief in sight: Ketotifen as a promising therapy for uremic pruritus in dialysis patients. Clin Med (Lond).* 2025;25(4 Suppl):100453. doi:10.1016/j.clinme.2025.100453

## Supplementary Material

Supplementary file 1. Pruritus in Dialysis Questionnaire [Pdf]

This file contains the questionnaire used for evaluating the severity, demographics, laboratory parameters and impact of uremic pruritus on sleep and quality of life among hemodialysis patients.

