## Supplementary file 1. Pruritus in Dialysis Questionnaire for "Ketotifen for Moderate-to-Severe Uremic Pruritus in Chronic Dialysis Patients: A Prospective Observational Study"

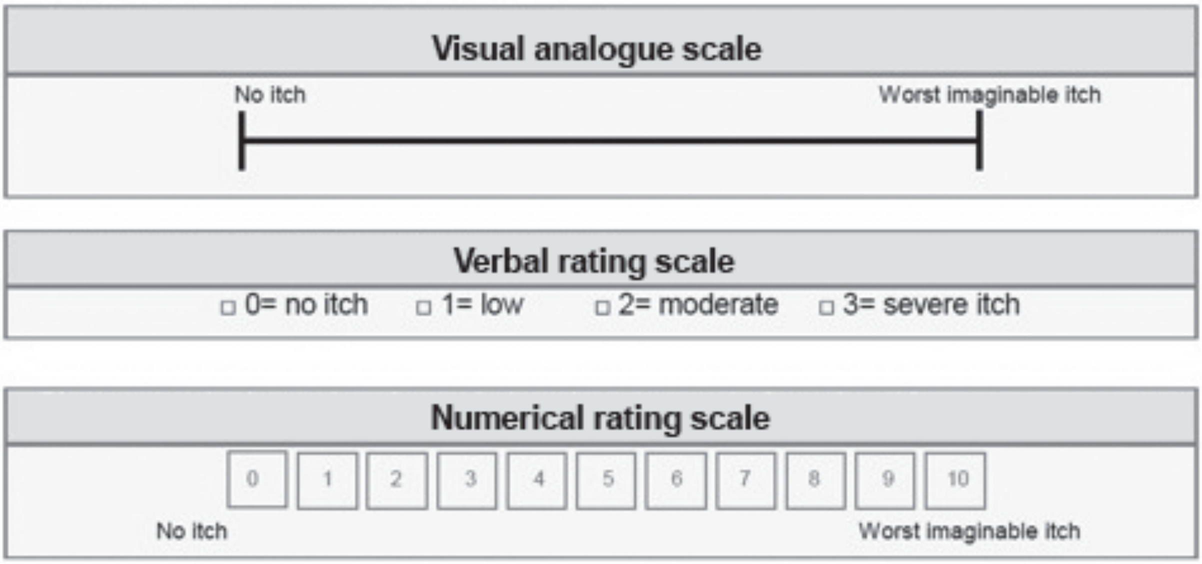


**Patient Information**

1. Name: __________________________________________
2. Age (in years): ___________
3. Hospital Number: ________________________________
4. Height (in cm): ___________
5. Weight (in kg): ___________

**Clinical Parameters**

6. Blood Pressure – Systolic (mmHg): ___________
7. Blood Pressure – Diastolic (mmHg): ___________

**Clinical Signs**

8. Presence of Edema: ☐ Yes (1)   ☐ No (2)
9. Presence of Skin Papules: ☐ Yes (1)   ☐ No (2)
10. Shiny Nails: ☐ Yes (1)   ☐ No (2)

**Medical History**

11. Basic Renal Disease (tick one):
    ☐ Diabetes (1)   ☐ Hypertension (2)   ☐ Chronic Glomerulonephritis (CGN)(3)
    ☐ Others (4) : ___________
12. Duration of Dialysis (in months): ___________
13. Total Number of Dialysis Sessions till Date: ___________

**Laboratory Results**

14. Hemoglobin (g/dL): ___________
15. Total Count (cells/mm³): ___________
16. ESR (mm/hr): ___________
17. Pre-Dialysis Blood Urea (mg/dL): ___________
18. Pre-Dialysis Serum Creatinine (mg/dL): ___________
19. Blood Sugar (mg/dL): ___________
20. HbA1C (%): ___________
21. Serum Calcium (mg/dL): ___________
22. Serum Phosphorus (mg/dL): ___________
23. Serum Uric Acid (mg/dL): ___________

**Treatment Details**

24. Number of Tablets per Day of Calcium Binders:
    - Calcium Carbonate: ___________
    - Sevelamer: ___________
    - Others (specify): ___________________________
25. Drugs for Itching: ☐ Yes (1) ☐ No (2)

**Symptom Management**

26. Sleep Disturbance due to Itching: ☐ Yes (1) ☐ No (2)

---

### Informed Consent

I, ______________________________________ (Name of Participant),
voluntarily agree to participate in this study. I understand that my medical information will be kept strictly confidential and used solely for research purposes. I have the right to withdraw at any point without affecting my treatment.

**Signature of Participant:** ____________________________ **Date:** ___________
**Signature of Investigator:** ____________________________ **Date:** ___________
